# Detecting early loss of kidney function in a Sri Lankan cohort study of working age adults

**DOI:** 10.64898/2026.08.18.26360601

**Authors:** Pubudu Chulasiri, Charlotte E Rutter, Nalika Gunawardena, SC Wickramasinghe, Rimaza Niyas, Neil Pearce, Ben Caplin, Thilanga Ruwanpathirana

## Abstract

**Objectives:** Chronic kidney disease of undetermined cause (CKDu) is a form of kidney disease not associated with traditional risk factors such as hypertension, diabetes or heavy proteinuria. 11.2% of men and 3.7% of women demonstrated low eGFR (a surrogate for CKDu) in the absence of these risk factors in a 2017 cross-sectional population-representative survey of adults in North Central Province, Sri Lanka. We therefore established a longitudinal cohort to track changes in kidney function over time and to identify environmental risk factors for developing poor kidney health.

**Methods:** This was a 6-year study of adults aged 20-60 years conducted in Puhudivula, Anuradhapura district. Exclusions included evidence of diabetes, hypertension or pre-existing CKD. We fitted hidden Markov models (HMMs) to estimate underlying state of kidney health and examine risk factors associated with departure from a healthy state.

**Results:** We identified four kidney health trajectories in the population (n=425): always healthy (74%); unhealthy throughout (5%); transition from health to unhealthy (10%); and reversion from unhealthy to healthy (11%). Using smokeless tobacco, including betel quid, was associated with being in an unhealthy category (2.29 [1.17, 4.49]). Lagged exposure to smoking (2.26 [1.25, 4.10]), smokeless tobacco (1.98 [1.13, 3.48]) and weedkiller (1.72 [1.15, 2.59]) were associated with the point of transition to an unhealthy state.

**Conclusions:** Almost a quarter of working age adults in this population demonstrated eGFR changes consistent with poor kidney health. Smokeless tobacco use was associated with both pre-existing evidence of poor kidney health and transitioning to the unhealthy category.

## Introduction

Chronic kidney disease of undetermined cause (CKDu) is a form of kidney disease that is not associated with traditional risk factors such as hypertension, diabetes or heavy proteinuria. It represents a considerable public health problem among some rural farming communities in Sri Lanka.

A cross-sectional representative population survey of rural Sri Lankan adults, (following the DEGREE protocol [1]), found prevalences of low eGFR (<60 ml/min per 1.73 m^2^) in the absence of these risk factors (a surrogate marker of CKDu) of 11.2% in men and 3.7% in women [2].

Such prevalence surveys can identify areas where CKDu is epidemic. However, to identify the causes of these population patterns, longitudinal studies are required in which initially healthy participants are followed over time, to identify which risk factors predict decline in eGFR [3]. We have previously conducted such a longitudinal study in Nicaragua. This used two different approaches to analyse eGFR trajectories. The first used growth mixture models (GMM) and identified three subgroups of participants, including one group (about 10% of men and 3% of women) with rapid decline in eGFR over the follow-up period. Factors associated with this rapid decline, and therefore likely exacerbating factors of disease, were men working in agriculture or outdoor work, along with lack of shade during work breaks [4]. The second approach used hidden Markov models (HMMs) to assess risk factors for departure from a state of kidney health. This approach identified that self-reported weight loss, nausea, vomiting and cramps, higher cumulative time in sugarcane work, excess occupational sun exposure, and NSAID use were each associated with the first evidence for loss of eGFR (a surrogate for point of disease onset) [5].

We have now conducted a similar longitudinal study in Sri Lanka using the same protocol [6]. This involved a cohort of working age adults (age 18-60 years) in rural Sri Lanka in which we tracked changes in kidney function over time and attempted to identify risk factors for disease onset, i.e., initial loss of kidney function.

## Methods

### Cohort

This was a longitudinal, community-based study of adults aged 18-60 years in the Puhudivula area of the Anuradhapura district. This area was selected for having the lowest mean eGFR of the 5 areas in a previous cross-sectional survey conducted in 2017 [2].

Participants were recruited in 2018 from participants in the previous cross-sectional survey; there was additional community recruitment using random sampling stratified by sex. Participants with self-reported CKD or eGFR <60 ml/min per 1.73 m^2^ at recruitment were excluded from joining. Participants were also excluded for self-reported hypertension or diabetes (with evidence of medical records), being on medication or treatment for hypertension or diabetes, current (at time of survey) blood pressure measured at or above 140 systolic and/or 90 diastolic, fasting plasma glucose >126 mg/dl. Additionally, pregnant women and those undergoing cancer treatment were excluded.

Follow-up visits took place in 2018, 2019, 2021, and 2023; thus, the maximum length of follow-up including the initial cross-sectional survey, was 6 years. At each visit, self-reported pregnant women were excluded from that study phase.

### Procedures

Similar to the previous study in Nicaragua [6], questionnaire data, clinical measurements and biosamples were collected at baseline and then at each follow-up. Participants were asked to respond to questions on demography, occupational history and current job, lifestyle factors (including diet, alcohol intake and tobacco use), medication use, exposure to sunlight and chemicals, and drinking water sources. Body weight was measured with minimal clothes using electronic scales and height using a portable stadiometer. Body composition was measured using a TANITA SC-240MA body composition analyser. Blood pressure was measured in a sitting position using an Omron HEM-7270 calibrated digital sphygmomanometer after five minutes of quiet seated rest. A mean of three measurements was recorded. Participants were asked to attend fasted, first thing in the morning (prior to work) in an attempt to reduce within- and between person variation in serum creatinine. The 2017 data collection from the cross-sectional study contained a subset of these measures. Full details of the data available at each time point is in the supplement (Supplementary Table S1).

### Biochemical methods

Serum creatinine was measured in a single batch using quality control referenced to international standards (for creatinine: isotope dilution mass-spectrometry quantified National Institute of Standards and Technology Standard Reference Material 967). The outcome measure was eGFR, calculated using the creatinine-based CKD-EPI 2021 equation [7].

### Statistical methods

Participants with fewer than 3 time-points of eGFR measurement were excluded. Time was defined as years since baseline (first survey including 2017 cross-sectional survey). Baseline characteristics and longitudinal eGFR measures were tabulated and the raw eGFR trajectories were plotted by sex.

We fitted hidden Markov models (HMMs) which estimate hidden states for each measurement (rather than each person’s measured trajectory), dependent only on the previous state, current eGFR (after adjustment for emission covariates), and any transition covariates. Each model was fitted 10 times on each of 1000 different initial values and the one with the best log-likelihood was automatically selected. Covariates were selected by comparing the Bayesian Information Criteria (BIC) for models including sex, baseline age and baseline age-squared (for non-linearity of age effect) as emission covariates. Based on our previous research [5], we fixed the distribution of the unhealthy state as ∼N(70,15^2^) to better reflect current medical knowledge and compared to a fully estimated model. We also considered models that used a 10% reduction in eGFR from baseline as a transition covariate.

From the final model we categorized each participant based on their changing states in the HMM. Those remaining in the healthy class throughout follow-up were categorized as “always healthy”, those always in the unhealthy class were labelled “always unhealthy”. Participants starting in the healthy category and transitioning to the unhealthy category and then staying there were classed as “healthy to unhealthy”. The final group, “reverted to healthy” are those who started in either class but at any point during the follow-up they transitioned from unhealthy to healthy.

We then estimated associations between potential risk factors and being “unhealthy” in the models. Firstly, we considered the baseline factors and whether they were associated with either of the person-level HMM groups that finished “unhealthy” compared to the “always healthy” group, using logistic regression models, adjusted for sex, age and education. Next, we considered a longitudinal model with a measure at each timepoint of the probability of departure from the healthy class. This is defined as the joint probability of being in an unhealthy state at the timepoint of interest and in a healthy state at the previous visit. This is only calculated for visits where the probability of being healthy in the previous visit was >50%. We used time varying risk factors and used the robust cluster sandwich estimator to account for the within person dependency. We used fractional logistic regression on this probabilistic outcome, adjusted for sex, baseline age, education and timepoint.

The time-varying exposures were measured at the same time as the outcome (i.e., every one or two years). We also estimated associations using a lagged exposure (i.e., using the exposure from the previous time-point) in case there was a delay between exposure and outcome.

We focused on the HMMs since these had given the most informative results in our previous analyses in Nicaragua. As noted above, we had also conducted GMM analyses in Nicaragua, and we have therefore performed the same analyses in Sri Lanka in order to enable a comparison. Thus, although we are focusing on the HMM results, we present the GMM results in an appendix. GMMs identify hidden groups of trajectories, with the eGFR of participants in each group coming from a normal distribution with random effects around a group mean eGFR. We used the LCMM R package which is widely used and has been validated against M plus using simulation studies [8]. We considered models adjusting for sex, age, age-squared and with sex as an effect modifier. Models were fitted for up to 4 hidden groups. To avoid local maximums, each model was fitted 1000 times with varying start values, and the one with the highest log-likelihood was selected. The BIC was used to select the overall best fitting model/number of groups. We estimated potential risk-factor associations with the probability of being in each group, using fractional logistic regression.

Analyses were conducted using R (HMM and GMM models) and Stata (descriptive, logistic and fractional logistic regression) software.

## Results

In the initial sample of 470 people, none were missing sex or age. We excluded 3 people with only 1 eGFR measurement and 42 with only 2 measurements, leaving a final analysis sample of 425 participants. (Supplementary Table S2)

In total there were 1827 valid eGFR measurements. They were fairly evenly spread across timepoints (between 359-419) except 2017 (283) which was smaller due to the additional recruitment in 2018. (Supplementary Table S3)

Just over half (55%) of the participants were female, the mean age was 36.7 years and the mean baseline eGFR was 116.9 ml/min per 1.73 m^2^ (Table 1). The trajectories appear similar in men and women, and it is difficult to visually identify different categories of eGFR trajectory (Figure 1). The mean eGFR was slightly lower for each subsequent time point (Supplementary Figure S1).

**Figure 1:**
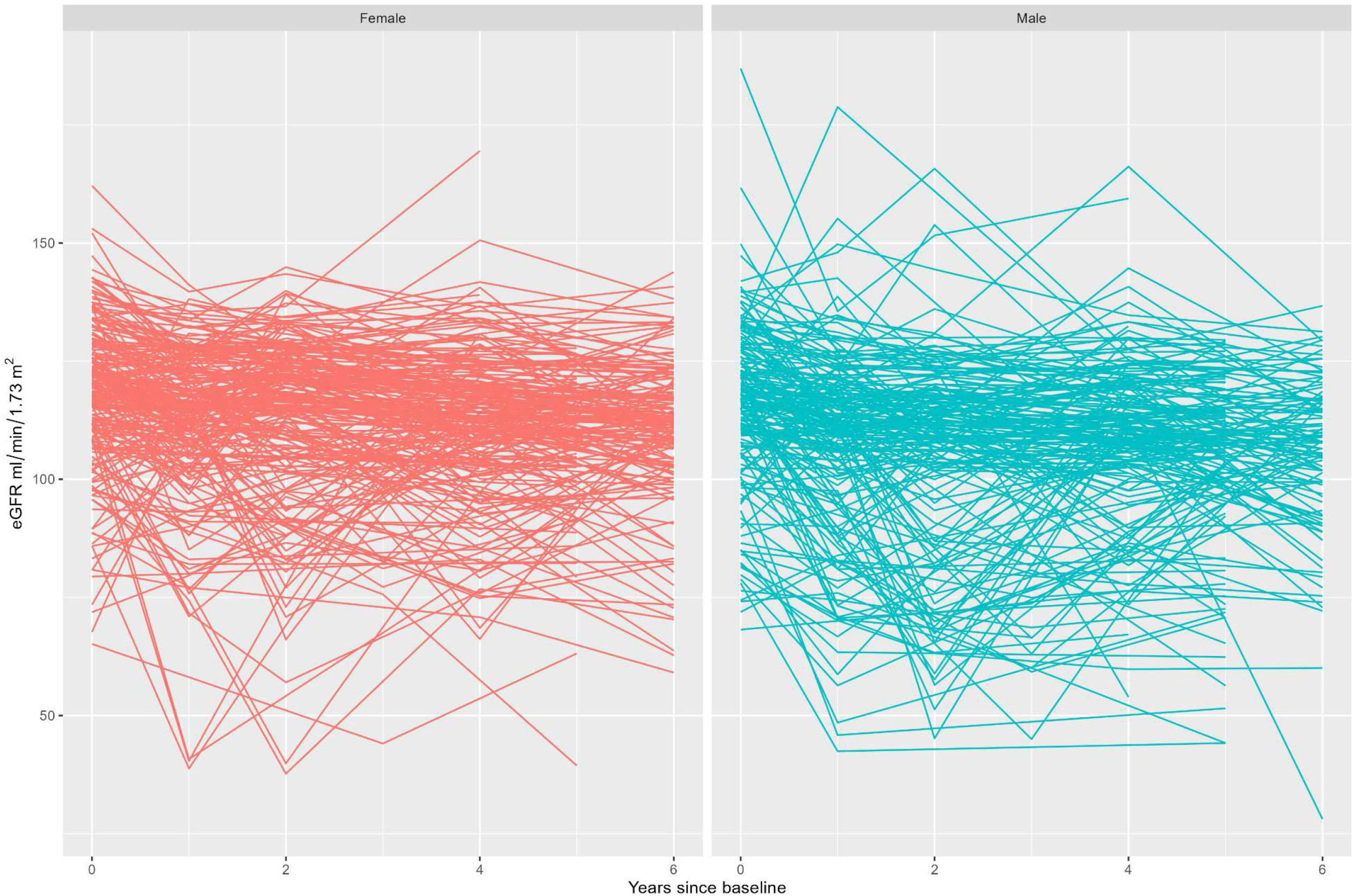
Observed trajectories of eGFR (mL/min/1.73m2) in Sri Lankan cohort by sex and years follow up (n=425)

**Table 1:** Baseline characteristics for those with at least 3 visits (n=425)

| Variable | Values | Total<sup>a</sup> | Female<sup>a</sup> | Male<sup>a</sup> |
| --- | --- | --- | --- | --- |
| Baseline year (first valid eGFR measure) | 2017 | 283 (67) | 167 (72) | 116 (60) |
|  | 2018 | 140 (33) | 64 (28) | 76 (40) |
|  | 2019 | 2 (0) | 1 (0) | 1 (1) |
| Available at time of first eGFR measure: |  |  |  |  |
| eGFR mL/min/1.7 m<sup>2</sup> | Mean (SD) | 116.9 (16.1) | 118.1 (15.4) | 115.3 (16.8) |
| Age in years | Mean (SD) | 36.7 (9.4) | 36.2 (9.3) | 37.3 (9.5) |
| Age group | 18-19 years | 11 (3) | 4 (2) | 7 (4) |
|  | 20-29 years | 96 (23) | 58 (25) | 38 (20) |
|  | 30-39 years | 127 (30) | 73 (32) | 54 (28) |
|  | 40-49 years | 170 (40) | 84 (36) | 86 (45) |
|  | 50-59 years | 21 (5) | 13 (6) | 8 (4) |
| BMI category (for South Asian populations) (n=424) | Underweight (<18.5 kg/m<sup>2</sup>) | 87 (21) | 39 (17) | 48 (25) |
|  | Normal weight (<23 kg/m<sup>2</sup>) | 172 (41) | 89 (38) | 83 (43) |
|  | Over weight (<27.5 kg/m<sup>2</sup>) | 107 (25) | 64 (28) | 43 (22) |
|  | Obese (>=27.5 kg/m<sup>2</sup>) | 58 (14) | 40 (17) | 18 (9) |
| Current Blood Pressure category | Normal blood pressure (<120/80) | 303 (71) | 171 (74) | 132 (68) |
|  | Elevated blood pressure (<130/80) | 53 (12) | 22 (9) | 31 (16) |
|  | HBP Stage 1 (<140/90) | 69 (16) | 39 (17) | 30 (16) |
| Fasting blood sugar | Normal (<100 mg/dL) | 217 (51) | 120 (52) | 97 (50) |
|  | Raised (<126 mg/dL) | 208 (49) | 112 (48) | 96 (50) |
| Available at 2018: |  |  |  |  |
| Education level | No school | 7 (2) | 4 (2) | 3 (2) |
|  | Grade passed | 192 (45) | 100 (43) | 92 (48) |
|  | O levels passed | 156 (37) | 85 (37) | 71 (37) |
|  | A levels passed | 57 (13) | 35 (15) | 22 (11) |
|  | Diploma | 7 (2) | 4 (2) | 3 (2) |
|  | University Degree | 6 (1) | 4 (2) | 2 (1) |
| Marital status | Married | 354 (83) | 200 (86) | 154 (80) |
|  | Unmarried | 57 (13) | 21 (9) | 36 (19) |
|  | Widowed | 7 (2) | 4 (2) | 3 (2) |
|  | Divorced | 7 (2) | 7 (3) | 0 (0) |
| Main occupation | Farming | 189 (45) | 109 (48) | 80 (42) |
|  | Housewife | 74 (18) | 74 (32) | 0 (0) |
|  | Government | 43 (10) | 3 (1) | 40 (21) |
|  | Private sector | 23 (5) | 8 (3) | 15 (8) |
|  | Shop keeper | 8 (2) | 5 (2) | 3 (2) |
|  | Self employed | 22 (5) | 7 (3) | 15 (8) |
|  | Labourer | 2 (0) | 0 (0) | 2 (1) |
|  | Student | 8 (2) | 4 (2) | 4 (2) |
|  | Vocational student | 7 (2) | 4 (2) | 3 (2) |
|  | Retired | 7 (2) | 1 (0) | 6 (3) |
|  | No job | 9 (2) | 5 (2) | 4 (2) |
|  | Other | 29 (7) | 9 (4) | 20 (10) |
| Duration of farming<br>(for those with main<br>occupation farming)<br>(n=184) | Up to 5 years | 35 (19) | 19 (18) | 16 (21) |
|  | Up to 10 years | 39 (21) | 19 (18) | 20 (25) |
|  | Up to 15 years | 26 (14) | 17 (16) | 9 (12) |
|  | Up to 20 years | 39 (21) | 27 (25) | 12 (15) |
|  | Up to 25 years | 21 (11) | 11 (10) | 10 (13) |
|  | Up to 30 years | 18 (10) | 10 (9) | 8 (10) |
|  | Over 30 years (max 45) | 6 (3) | 3 (3) | 3 (4) |
|  | Missing | 5 | 3 | 2 |
| Household income | 1=No income | 6 (1) | 5 (2) | 1 (1) |
|  | 2= <=10000 Rs | 37 (9) | 24 (10) | 13 (7) |
|  | 3= 10001-20000 Rs | 74 (17) | 40 (17) | 34 (18) |
|  | 4= 20001-30000 Rs | 106 (25) | 64 (28) | 42 (22) |
|  | 5= 30001-40000 Rs | 84 (20) | 49 (21) | 35 (18) |
|  | 6= 40001-50000 Rs | 53 (12) | 25 (11) | 28 (15) |
|  | 7= 50001-60000 Rs | 33 (8) | 17 (7) | 16 (8) |
|  | 8= >60000 Rs | 32 (8) | 8 (3) | 24 (12) |
| Smoking status<br>(n=424) | Never smoked | 307 (72) | 231 (100) | 76 (40) |
|  | Used to smoke | 33 (8) | 0 (0) | 33 (17) |
|  | Currently smoke | 84 (20) | 1 (0) | 83 (43) |
| Smokeless tobacco<br>status (n=421) | Never used | 330 (78) | 213 (92) | 117 (62) |
|  | Used to use | 14 (3) | 3 (1) | 11 (6) |
|  | Currently use | 77 (18) | 15 (6) | 62 (33) |
| Alcohol use<br>(n=409) | Daily | 3 (1) | 0 (0) | 3 (2) |
|  | 5-6 days a week | 3 (1) | 1 (0) | 2 (1) |
|  | 3-4 days a week | 9 (2) | 0 (0) | 9 (5) |
|  | 1-2 days a week | 13 (3) | 0 (0) | 13 (7) |
|  | 1-3 days a month | 19 (5) | 0 (0) | 19 (10) |
|  | Less than once a month | 17 (4) | 1 (0) | 16 (8) |
|  | Occasionally | 13 (3) | 1 (0) | 12 (6) |
|  | Consumed earlier but<br>not now | 57 (14) | 0 (0) | 57 (30) |
|  | Never consume(d)<br>alcohol | 275 (67) | 217 (98) | 58 (31) |
| Physical Activity level<br>(n=422) | Sedentary | 7 (2) | 6 (3) | 1 (1) |
|  | Minimum activity | 96 (23) | 70 (31) | 26 (13) |
|  | Medium activity | 187 (44) | 113 (49) | 74 (38) |
|  | Strenuous activity | 132 (31) | 40 (17) | 92 (48) |
| Excessive exposure to sunlight (n=422) |  | 231 (55) | 159 (69) | 72 (37) |
| Breaks from work when exposed to sunlight (n=418) | No sunlight | 227 (54) | 145 (63) | 82 (44) |
|  | Sunlight with breaks | 179 (43) | 81 (35) | 98 (52) |
|  | Sunlight no breaks | 12 (3) | 4 (2) | 8 (4) |
| Breaks from work in shade when exposed to sunlight (n=494) | No sunlight | 227 (56) | 145 (64) | 82 (46) |
|  | Breaks in shade | 121 (30) | 54 (24) | 67 (37) |
|  | No breaks in shade | 59 (15) | 28 (12) | 31 (17) |
| Any Reverse Osmosis water (n=422) |  | 284 (67) | 152 (66) | 132 (68) |
| Vegetarian diet (n=424) |  | 13 (3) | 9 (4) | 4 (2) |
| Fish in diet |  | 395 (93) | 215 (93) | 180 (93) |
| Meat in diet (n=424) |  | 369 (87) | 189 (81) | 180 (94) |
| Eggs in diet |  | 385 (91) | 207 (89) | 178 (92) |
| Exposed to fertilisers |  | 266 (63) | 117 (50) | 149 (77) |
| Exposed to weedkiller |  | 186 (44) | 53 (23) | 133 (69) |
| Exposed to roundup |  | 73 (17) | 12 (5) | 61 (32) |
| Exposed to pesticides |  | 124 (29) | 31 (13) | 93 (48) |

<sup>a</sup> showing n (%) for binary, n (column %) for categorical and mean (SD) for continuous variables; eGFR=estimated glomerular filtration rate; BMI=body mass index; HBP=high blood pressure

The best fitting HMM model (using the BIC) included sex, baseline age and age-squared plus the transition probability included a covariate for losing >10% eGFR. The models using fully estimated unhealthy states generally had lower BICs but we had decided a priori that using a fixed unhealthy distribution provided a more accurate definition of unhealthy kidneys, so this was selected as our final model (Supplementary Tables S4). We note that the unhealthy distribution from the best fitting fully estimated model was centred at 95 ml/min per 1.73 m^2^ (data not shown). In the final model the healthy eGFR distribution was estimated with mean 113.7 ml/min per 1.73 m^2^ (SD=9.73), and the unhealthy was fixed at mean 70 ml/min per 1.73 m^2^ (SD=15). Being male was associated with a 0.11 ml/min per 1.73 m^2^ lower eGFR across both states. (Supplementary Table S5)

Categorising the person-level trajectories from this final model, 316 people (74.4% of the cohort) were classed as healthy throughout, 19 (4.5%) were unhealthy throughout, 43 (10.1%) moved from healthy to unhealthy and remained unhealthy and a further 47 (11.1%) moved unhealthy and reverted back to healthy at some point. The proportion of women classed as healthy throughout was higher than the proportion of men (79% compared to 68%). (Figure 2 and Supplementary Table S5)

**Figure 2:**
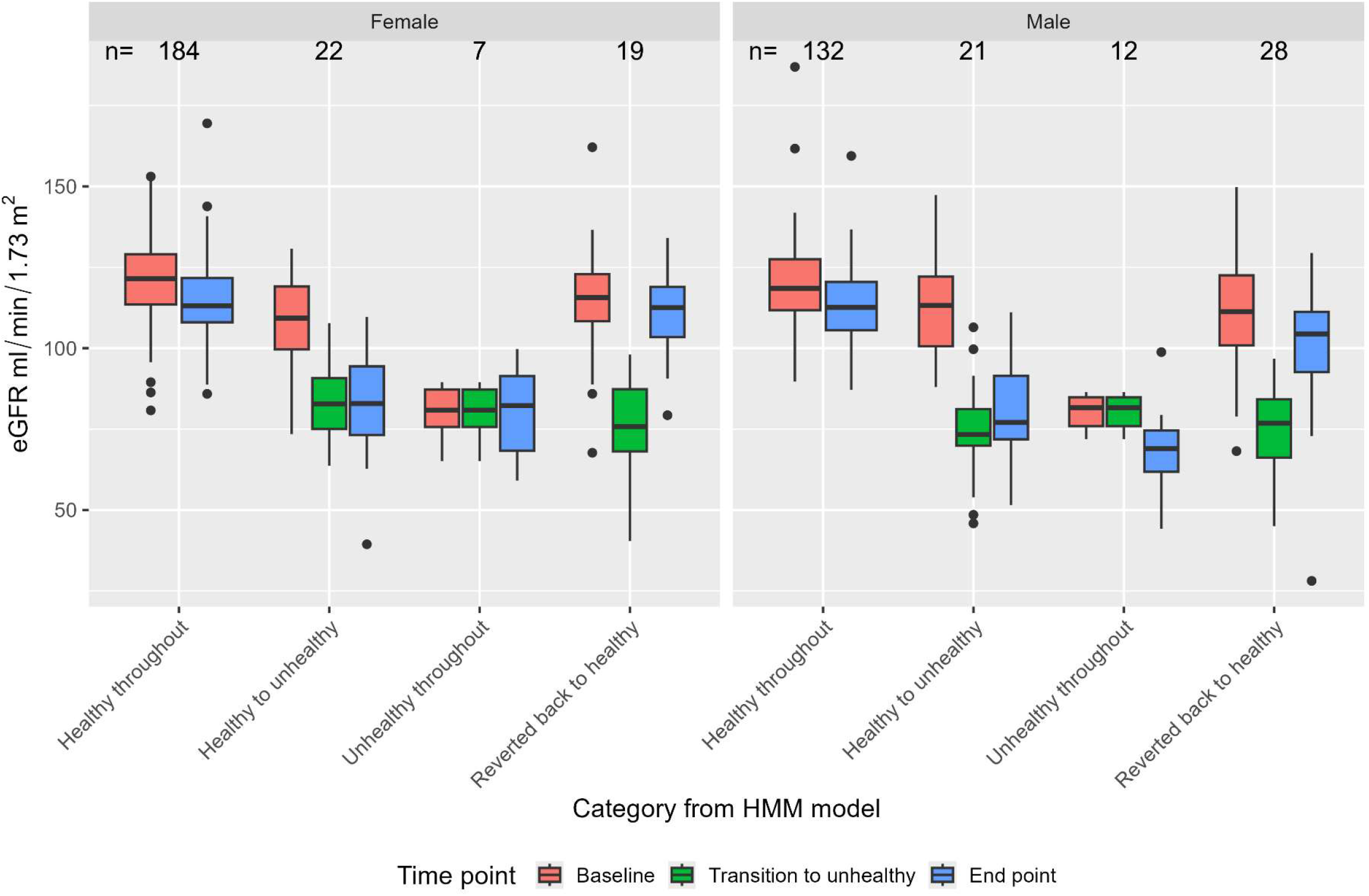
eGFR distributions at baseline, final visit, and when they first enter an unhealthy state, by sex and category of movement

The mean eGFR in the “healthy throughout” group is high at both baseline and end point, although does drop a little. In the “unhealthy throughout” group the eGFR stays low (and drops further for men). In the “healthy to unhealthy” group (the main group of interest) the eGFR is high at baseline and low at both the point of transition and at the end. The remaining group of people who were unhealthy at some point but then reverted to healthy showed high eGFR at baseline and end point, but low eGFR at the point of transition to unhealthy. These results are reassuring as they are similar to what would be expected of these groups. (Figure 2)

The only baseline characteristic that was associated with an increased risk of ever being in the unhealthy state, after adjusting for sex, age and education, was using smokeless tobacco (2.29 [1.17, 4.49]). (Table 2)

**Table 2:** Associations between baseline exposure and unhealthy groups^a^ in the Hidden Markov Model (HMM), adjusted for age, sex and education, using logistic regression

| Variable |  | HMM with fixed unhealthy distribution |  |  |
| --- | --- | --- | --- | --- |
|  |  | n | OR (95% CI) | p-value |
| Sex (Male) |  | 378 | 1.57 (0.91, 2.72) | 0.11 |
| Age (1-year increase) |  | 378 | 1.02 (0.99, 1.06) | 0.14 |
| Education level | No O levels | 378 | 1.06 (0.56, 1.98) | 0.87 |
|  | O levels |  | ref | NA |
|  | A levels and above |  | 1.17 (0.51, 2.69) | 0.71 |
| Main occupation is farming |  | 378 | 1.17 (0.64, 2.11) | 0.62 |
| Duration of farming | No farming | 373 | ref | NA |
|  | <10 years |  | 0.66 (0.27, 1.60) | 0.36 |
|  | <20 years |  | 1.96 (0.90, 4.31) | 0.09 |
|  | <30 years |  | 1.33 (0.51, 3.44) | 0.56 |
| Income group | Low (<=20000) | 378 | 1.49 (0.75, 2.96) | 0.25 |
|  | Medium |  | ref | NA |
|  | High (>40000) |  | 1.42 (0.72, 2.77) | 0.31 |
| BMI category | Underweight | 378 | 0.69 (0.31, 1.51) | 0.35 |
|  | Normal |  | ref | NA |
|  | Overweight |  | 0.81 (0.40, 1.64) | 0.55 |
|  | Obese |  | 1.08 (0.47, 2.45) | 0.86 |
| Painkiller tablets |  | 378 | 1.45 (0.77, 2.71) | 0.25 |
| Painkiller injections |  | 378 | 0.39 (0.05, 3.18) | 0.38 |
| Current smoker |  | 377 | 1.75 (0.81, 3.77) | 0.15 |
| Current smokeless tobacco user |  | 377 | 2.29 (1.17, 4.49) | 0.02 |
| Drinks Alcohol 2+ times a week |  | 364 | 1.84 (0.44, 7.60) | 0.40 |
| Very physically active |  | 375 | 1.10 (0.59, 2.06) | 0.76 |
| Excessive exposure to sunlight |  | 375 | 0.76 (0.42, 1.35) | 0.35 |
| Breaks from work when exposed | No sunlight | 371 | ref | NA |
|  | Sunlight with breaks |  | 0.84 (0.47, 1.50) | 0.55 |
|  | Sunlight no breaks |  | 0.37 (0.05, 3.07) | 0.36 |
| Breaks from work in shade when exposed | No sunlight | 360 | ref | NA |
|  | Breaks in shade |  | 0.87 (0.45, 1.67) | 0.67 |
|  | No breaks in shade |  | 0.90 (0.40, 2.06) | 0.81 |
| Reverse Osmosis water use |  | 375 | 1.70 (0.89, 3.25) | 0.11 |
| Vegetarian diet |  | 377 | 1.09 (0.23, 5.19) | 0.91 |
| Fish in diet |  | 378 | 0.59 (0.24, 1.45) | 0.26 |
| Meat in diet |  | 377 | 1.96 (0.73, 5.26) | 0.18 |
| Eggs in diet |  | 378 | 0.88 (0.36, 2.12) | 0.78 |
| Fertilizer exposure |  | 378 | 1.05 (0.56, 1.97) | 0.89 |
| Weedkiller exposure |  | 378 | 1.28 (0.65, 2.52) | 0.47 |
| Roundup exposure |  | 378 | 1.16 (0.56, 2.42) | 0.69 |
| Pesticide exposure |  | 378 | 1.37 (0.72, 2.63) | 0.34 |

<sup>a</sup> healthy to unhealthy or always unhealthy groups compared to the always healthy group; OR, odds ratio; 95% CI, 95% confidence interval; BMI, Body Mass Index.

For time-varying exposures and the probability of departure from the healthy distribution of the HMM, we did not initially see any evidence of associations (Table 3). Once we used a lagged exposure (i.e. from the previous time point) we saw evidence that current smoking (2.26, [1.25, 4.10]), using smokeless tobacco (1.98, [1.13, 3.48]) and reported exposure to weedkiller (1.72, [1.15, 2.59]), even though the sample size was reduced as the first time point could not be used (Table 3).

**Table 3:** Associations between time varying exposures and the probability of departure from the healthy eGFR distribution in the Hidden Markov Model with fixed unhealthy distribution, using fractional logistic regression with the cluster sandwich estimator to account for within person dependency and adjusted for sex, baseline age, education and timepoint

| Time-varying exposure | Risk factor associations<sup>a</sup> |  |  | Lagged risk factor associations<sup>b</sup> |  |  |
| --- | --- | --- | --- | --- | --- | --- |
|  | n | OR (95% CI) | p-value | n | OR (95% CI) | p-value |
| Main occupation farming | 1179 | 1.15 (0.76, 1.76) | 0.50 | 889 | 1.27 (0.79, 2.02) | 0.32 |
| Weight loss >2.5kg | 1202 | 0.76 (0.42, 1.40) | 0.39 | 858 | 1.28 (0.71, 2.30) | 0.41 |
| Painkiller tablets | 1199 | 1.26 (0.79, 2.02) | 0.34 | 914 | 0.87 (0.49, 1.53) | 0.63 |
| Painkiller injections | 1199 | 0.55 (0.14, 2.15) | 0.39 | 914 | 0.68 (0.19, 2.42) | 0.56 |
| Current smoker | 1198 | 0.72 (0.41, 1.26) | 0.25 | 913 | 2.26 (1.25, 4.10) | 0.007 |
| Current smokeless tobacco user | 1197 | 1.09 (0.67, 1.77) | 0.74 | 912 | 1.98 (1.13, 3.48) | 0.02 |
| Drink Alcohol 2+ times a week | 1165 | 0.44 (0.12, 1.68) | 0.23 | 885 | 2.24 (0.86, 5.80) | 0.10 |
| Strenuous activity | 1197 | 0.77 (0.49, 1.22) | 0.26 | 911 | 1.19 (0.75, 1.88) | 0.47 |
| Excessive exposure to sunlight | 1197 | 0.88 (0.59, 1.29) | 0.51 | 911 | 1.04 (0.68, 1.60) | 0.86 |
| Sun exposure with no breaks in shade | 1140 | 0.69 (0.25, 1.85) | 0.46 | 870 | 0.96 (0.48, 1.95) | 0.92 |
| Use of RO water | 1191 | 1.09 (0.66, 1.80) | 0.74 | 905 | 1.26 (0.78, 2.05) | 0.35 |
| Vegetarian | 1198 | 0.79 (0.27, 2.72) | 0.66 | 913 | 0.74 (0.26, 2.16) | 0.59 |
| Fertiliser exposure | 1199 | 0.75 (0.50, 1.13) | 0.17 | 914 | 1.36 (0.87, 2.12) | 0.18 |
| Weedkiller exposure | 1199 | 0.85 (0.54, 1.33) | 0.48 | 914 | 1.72 (1.15, 2.59) | 0.009 |
| Roundup exposure | 1194 | 0.66 (0.34, 1.29) | 0.22 | 913 | 1.38 (0.74, 2.56) | 0.31 |
| Pesticide/insecticide exposure | 1199 | 0.77 (0.47, 1.24) | 0.28 | 914 | 1.16 (0.71, 1.91) | 0.55 |

<sup>a</sup> Exposure since last time-point; <sup>b</sup> Exposure between previous 2 time-points; OR, odds ratio; 95% CI, 95% confidence interval; RO, Reverse Osmosis

With the GMMs, the best fitting model (using BIC) was the 2-class model adjusting for sex and baseline age (Supplementary Table S6). This model identified a group with higher, more stable eGFR (78.6% of the cohort) and a group with lower more erratic eGFR (21.4%) but did not identify a group with obvious declining eGFR. (Supplementary Figure S2 and Table S7). The 3-class models had a worse fit and still did not identify a group with obviously declining eGFR (data not shown). The groups identified using HMM and those using GMM are compared in Supplementary Table S8 showing there was reasonable alignment between them.

Risk factor analyses using the classes from the 2-state GMM model showed that men were 50% more likely to be in the lower erratic group than women (OR=1.50, 95% CI [1.07, 2.10]). There was only very weak evidence that people using smokeless tobacco were more likely to be in the lower erratic group (1.54 [0.96, 2.49]). (Supplementary Table S9)

## Discussion

This is the first longitudinal study in Sri Lanka to track eGFR over time in the apparently healthy at-risk rural population from an area with high prevalence of CKDu. The cohort has been followed up for 6 years and is evenly split by sex and occupation (farming/non-farming), with about half of both sexes working in farming. In the area of the study, the main crop is rice (paddy), but other fruit and vegetables are also cultivated.

The HMM with a fixed unhealthy state (70 ml/min per 1.73 m^2^) was similar to the model used in Nicaragua and ensured that the unhealthy definition was closer to the clinical definition of CKD. In this model we estimated that 10.1% of the cohort (men 10.9% and women 9.5%) became and stayed unhealthy during follow up with a slightly larger proportion (11.1%) becoming unhealthy but reverting back to healthy (men 14.5% and women 8.2%).

By comparison, the HMM in the Nicaraguan cohort showed that 15% of men and 5% of women moved from healthy to unhealthy during the follow up but that only 3% overall reverted to healthy from unhealthy [5].

We identified two groups of kidney function trajectories using GMMs. The first (79%) was a high and stable eGFR group. The second (21%) was a lower and more erratic eGFR group. There was a much higher proportion of men in the erratic group than women (26% compared to 17%).

These findings demonstrate a different pattern of disease than that shown in the similar analysis of the Nicaraguan cohort which identified a “rapid eGFR decline” group [4].

The two different types of models (HMM and GMM) each showed similar patterns, albeit in different ways. We identified an erratic group using GMM and not a rapid declining group, so it is perhaps not surprising that the HMM showed 11% reverting back to healthy. It should be emphasized that “reverting to healthy” is not a clinical descriptor but is a statistical descriptor using an estimate of the best fitting state distribution.

Thus, we see a difference in eGFR trajectories here compared to Nicaragua. This is consistent with other speculation [9] that CKDu may have different characteristics, and also possibly different causes, in Central America (including Nicaragua) and South Asia (South India and Sri Lanka). For example, the observed patterns in Sri Lanka may be due to exposure to different CKD-exacerbating factors. That is, whether or not the primary cause of the disease is the same between the two regions, the absence of the high levels of occupational heat exposure that occurs in Central America may explain why no equivalent of the ‘rapid decline’ group is seen in Sri Lanka.

We identified that using smokeless tobacco was associated with increased risk of ever being classed as unhealthy in the HMM and very weak evidence of being in the lower erratic eGFR class in the GMM. The GMM also showed evidence that being male was an increased risk of being in the lower erratic class (there was no evidence in the HMM but the direction of the effect estimate was consistent). This similarity between quite different statistical methods lends credence to the findings. In Sri Lanka the most common form of smokeless tobacco used is betel quid, which mixes tobacco with areca (betel) nut and slaked lime, wrapped in a betel leaf and is chewed. Areca nuts are a known carcinogen [10] and have been shown to be associated with CKD [11].

An advantage of the HMM is that we were able to include time-varying covariates, with the aim of identifying factors associated with the time of disease onset (or at least the start of a decline in eGFR). We saw harmful associations in the time varying analysis using a lagged exposure of both smoking (2.26 [1.25, 4.10]), use of smokeless tobacco (1.98 [1.13,3.48]) and exposure to weedkiller (1.72 [1.15, 2.59]).

These associations could be causally linked, either to actual disease onset or to progression of kidney dysfunction in those already with the disease. But it is also possible that these associations were due to confounding from other unmeasured factors.

The main strengths of this study are that this is the first and only community cohort study of kidney function in Sri Lanka, and the analysis methods allow for variability in an individual’s baseline kidney function (compared to using one cut-off point of eGFR) and focus on the changes over time. Additionally, there was a large amount of data collected on potential associated lifestyle and occupational factors.

Limitations include that the risk factor information was largely collected by questionnaire, and there were no biological measurements of risk factors in the current analyses.

### Conclusion

We found that around a quarter of working age adults in rural Anuradhapura district had eGFR changes consistent with either new-onset or pre-existing poor kidney health but did not show a rapid decline in kidney function. This is positive news that may imply exacerbating factors are not as severe as in other areas, although underlying CKDu will still result in loss of kidney function and high mortality but over a longer time-period. We identified that those who use smokeless tobacco were at increased risk of poor kidney health.

## Ethics Approval

Ethical approval (EC-25-170) was granted by the Ethics Review Committee, Faculty of Medicine, University of Colombo, Sri Lanka.

## Acknowledgments

We would like to thank the participants and local communities for enabling this study to be conducted.

## Data Availability

Information on data availability can be obtained by contacting Dr Thilanga Ruwanpathirana, Ministry of Health, Sri Lanka.

## Author Contribution

NP and BC conceived the idea of the study; NP, BC, PC, NG and TR designed the study; PC, NG, TR collected the data; CER analysed the data; CER, NP, and BC wrote the first draft of the manuscript; all other authors contributed to revisions of the manuscript.

