## Supplementary for "Detecting early loss of kidney function in a Sri Lankan cohort study of working age adults"

Supplementary material

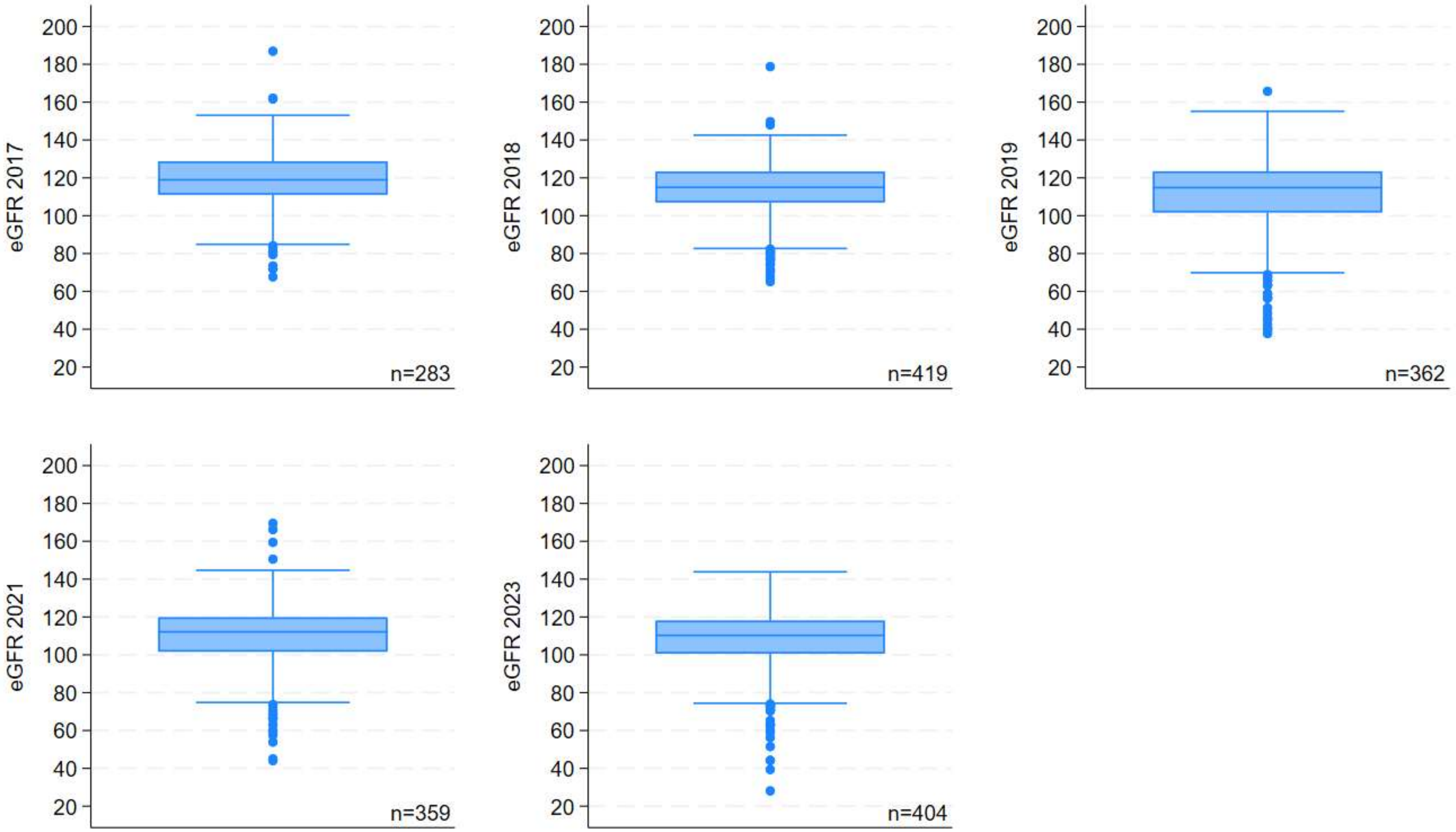


Figure S1: Box plots of eGFR (mL/min/1.73m$^2$) distributions by visit year for participants with 3 or more measures

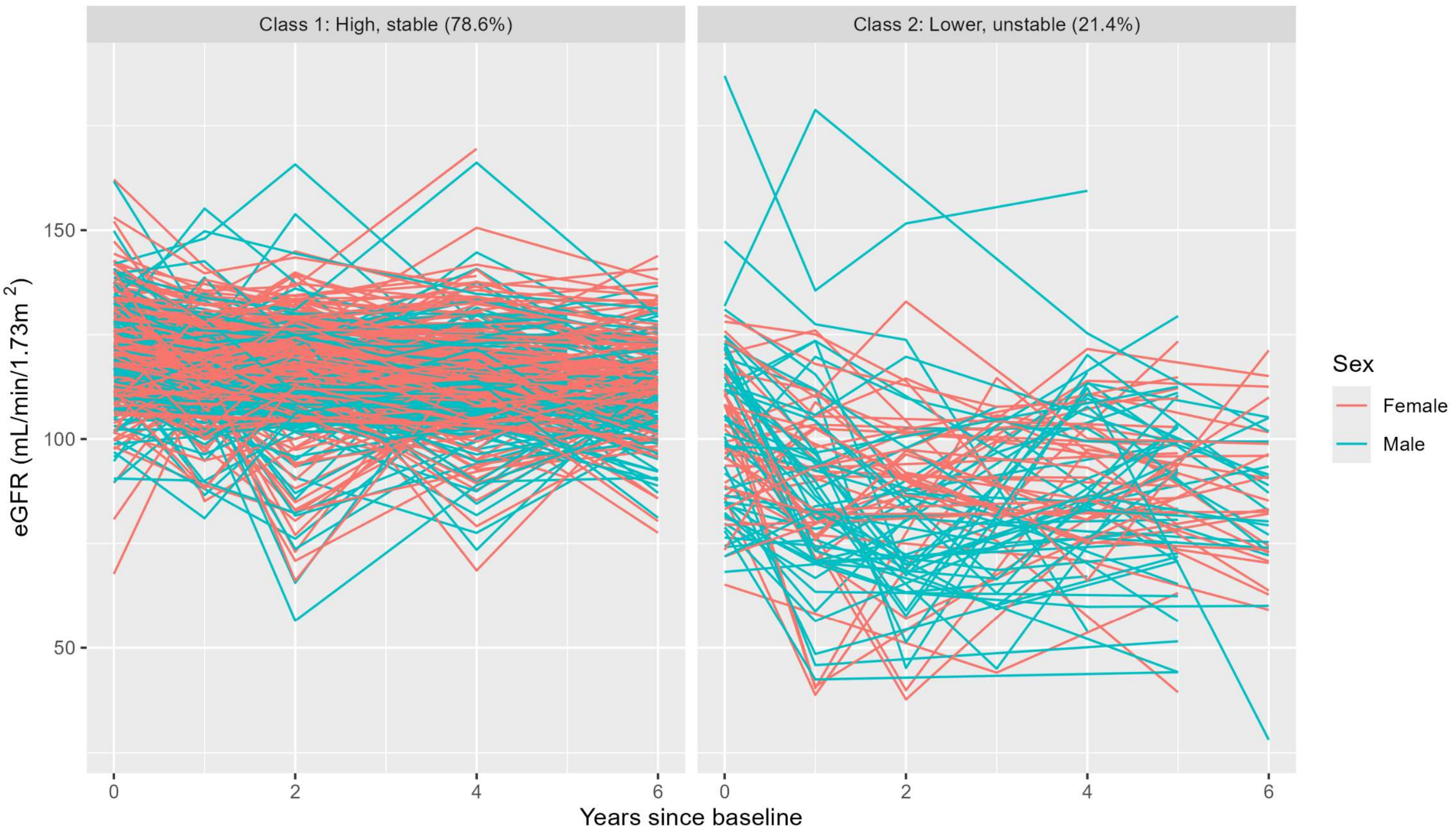


Figure S2: Trajectories of eGFR (mL/min/1.73m$^2$) by class and sex from 2 class GMM model with sex and age adjustment (n=425)

Table S1: Questionnaire and variable definitions by time point

| Variables | Definition | Time points |
|---|---|---|
| eGFR (mL/min/1.7 $m^2$) | CKD-EPI 2021 calculated using serum creatinine | All |
| Sex | Female/Male | All |
| Age (years) | | All |
| Weight (kg) | | All |
| Body Mass Index | Weight (kg) / Height $(m)^2$ | All (Height only measured in 2018 but should be stable) |
| Measured Blood Pressure | Average systolic and diastolic of 3 readings | All |
| Fasting blood sugar | Capillary RBS | 2017-2019 (Random Blood Sugar measured 2021-2023) |
| Education level | What is the highest level of education you have completed? No school, grade passed, O levels passed, A levels passed, diploma, university degree | 2018-2023 |
| Marital status | What is your marital status? Married, unmarried, divorced, widowed | All |
| Main occupation | If you are working what is your main occupation? | 2017 |
| | Currently your main occupation is farming?<br>If you did not have farming as your main occupation after the last survey, what would have been your main occupation? | 2018-2023 |
| Duration of farming | (For those with main occupation farming) What duration have you engaged in the following farming/industries as the main or part time occupation? | 2018-2023 |
| Household income | What is your family's monthly family income? (8 groups) | 2018-2023 |
| Smoking status | Now I am going to ask you some questions about tobacco use for last one year.<br>Did you smoke any tobacco products, such as cigarettes or Beedi? | 2017 |
| | Have you smoked cigarettes or beedi after the last survey?<br>Do you currently smoke? | 2018-2023 |
| Smokeless tobacco status | Now I am going to ask you some questions about smokeless tobacco use for last one year [such as chewing tobacco, betel with tobacco, babul, snuff ]<br>In the past one year, did you use smokeless tobacco products? | 2017 |

| | | |
|---|---|---|
| | I'm going to ask you a few more questions about tobacco use after the last survey. (Tobacco, tobacco with betel nut, babulle, tobacco smoke)<br>Did you use the tobacco-containing substance mentioned above after the last survey?<br>Are you currently using the tobacco-containing substance mentioned above? | 2018-2023 |
| Alcohol use | Now I am going to ask you some questions about alcohol use (such as arrack, kasippu, toddy, beer, spirits or wine). During the past 12 months, how frequently have you had at least one standard alcoholic drink?<br>Daily, 5-6 days per week, 3-4 days per week, 1-2 days per week, 1-3 days per month, less than once a month, not at all. | 2017 |
| | I'm going to ask you a few questions about alcohol after the last survey. (Arrack, kassippu. Toddy, beer, spirits or wine). Nowadays the frequency of your drinking can be better described:<br>Daily, 5-6 days per week, 3-4 days per week, 1-2 days per week, 1-3 days per month, once a month, less than once a month, occasionally when going to a party, drank in the past but does not drink at present, never drink alcohol. | 2018-2023 |
| Painkiller use | Have you taken the following medications after the last survey?<br>Painkiller capsules like Panadol / panadine / paracetamol<br><br>Pain killer injections like Diclofenac / celecox | 2018-2023 |
| Physical Activity level | What is the following description of your physical activity, as a whole, taken after the last survey? Here employment, transportation, and physical activity for sports should all be considered.<br>1. Excessive physical activity (e.g. heavy lifting / lifting / land digging / construction or sports activities that increase heart activity or increase respiration rate),<br>2. Normal physical activity (eg, slightly increased heart rate or increased respiratory rate, walking or light weight lifting),<br>3. Minor physical activity (e.g. minimal physical activity),<br>4. No physical activity (sitting or sleeping) | 2018-2023 |
| Excessive exposure to sunlight | Have you been exposed to too much sunlight in the last year? | 2018-2023 |

| | | |
|---|---|---|
| Breaks from sunlight | Did you get breaks while working while being exposed to excessive sunlight during the past year? | 2018-2023 |
| Inside breaks from sunlight | During the past year, have you been exposed to too much sunlight and had any shade during your breaks at work? | 2018-2023 |
| Any Reverse Osmosis water for drinking | Please state your main water source used for drinking and cooking purposes for last one year/since last survey. You may have multiple water sources.<br><br>From the RO water sources in the village, From the RO water source in his home or private place, Filtered water for sale, Sealed water bottles purchased at the store, From a deep well, From the cultivation well, From tube well, Piped Water from water board/by road water By the water sources in the tanks, By community water sources, other. | 2018-2023 |
| Diet | What type of food are you currently consuming? | |
| Vegetarian diet | Strictly vegetarian (yes/no) | 2018-2023 |
| Fish in diet | Fish (yes/no) | 2018-2023 |
| Meat in diet | Meat (yes/no) | 2018-2023 |
| Eggs in diet | Eggs (yes/no) | 2018-2023 |
| Chemical exposure | Did you apply / mix the following ingredients last year?<br>Were you exposed to chemical fertilizers/weedicides/ pesticides for last one year? The exposure can be by way of using them in farming or for other purposes or handling them to store | |
| Exposed to fertilisers | Chemical fertilisers | 2017 |
| | Blue cubes, red powder, flower fertilizer,<br>Urea, TDM, STP, mud fertilizer, MOP 3,4 DPA | 2018-2023 |
| Exposed to weedkiller | Weedicides | 2017 |
| | Satinil 600 CE, M 50, Nomini, Gulliver, Agroxone, MCP 60, Gramxone, Headenol, PowerMate | 2018-2023 |
| Exposed to roundup | Round up or some other similar object | 2018-2023 |
| Exposed to pesticides | Pesticides | 2017 |
| | B.P.M.C., Curator, Mospila, Elson, Admeyer, Soro, Marshall 20, Korojan, Dithian, Calcon, Mig 18 | 2018-2023 |

Table S2: Number of visits per participant (with valid eGFR measure)

| Number of visits | Overall<br>n (col %) | Female<br>n (col %) | Male<br>n (col %) |
|---|---|---|---|
| 1 (excluded) | 3 (0) | 1 (0) | 2 (1) |
| 2 (excluded) | 42 (7) | 11 (4) | 31 (14) |
| 3 | 104 (17) | 49 (18) | 55 (24) |
| 4 | 90 (15) | 38 (14) | 52 (23) |
| 5 | 231 (38) | 145 (52) | 86 (38) |
| Total included | 425 | 232 | 193 |

Table S3: Number of participants per visit (from those with at least 3 eGFR measures)

| Characteristic | Year of visit | Total | Female | Male |
|---|---|---|---|---|
| Number of eGFR measures (row %) | 2017 | 283 | 167 (59) | 116 (41) |
| | 2018 | 419 | 230 (55) | 189 (45) |
| | 2019 | 362 | 201 (56) | 161 (44) |
| | 2021 | 359 | 202 (56) | 157 (44) |
| | 2023 | 404 | 224 (55) | 180 (45) |
| | All | 1827 | 1024 (56) | 803 (44) |

Table S4: Comparison of Bayesian Information Criteria for each Hidden Markov Model, all with eGFR as the main outcome (n=1827)

| Emission covariates (affect eGFR along with hidden state) | No transition covariates; fixed unhealthy state ~N(70,15$^2$) | >10% loss of eGFR; fixed unhealthy state ~N(70,15$^2$) | No transition covariates; estimated states | >10% loss of eGFR; estimated states |
|---|---|---|---|---|
| None | 15295.0 | 15288.0 | 15184.4 | 15179.3 |
| Sex | 15294.9 | 15288.4 | 15174.2 | 15168.2 |
| Sex and age | 14590.4 | 14583.0 | 14367.8 | 14369.9 |
| Sex, age and age-squared | 14581.0 | 14573.2 | 14367.2 | 14369.1 |

Table S5: Hidden Markov Model results from final model with unhealthy distribution fixed at ~N(70,$15^2$), sex and age emission covariates and 10% drop in eGFR as a transition covariate

| Person-level category | Distribution n (%) | | | Healthy state, mean eGFR ml/min per 1.73 $m^2$ (SD) | | Unhealthy state, mean eGFR ml/min per 1.73 $m^2$ (SD) | |
|---|---|---|---|---|---|---|---|
| | All | Female | Male | Female | Male | Female | Male |
| Stayed healthy | 316 (74.4) | 184 (79.3) | 132 (68.4) | 113.69 (9.73) | 113.58 (9.73) | 70.00 (15) | 69.88 (15) |
| Healthy to unhealthy | 43 (10.1) | 22 (9.5) | 21 (10.9) | | | | |
| Stayed unhealthy | 19 (4.5) | 7 (3.0) | 12 (6.2) | | | | |
| Reverted to healthy | 47 (11.1) | 19 (8.2) | 28 (14.5) | | | | |

Table S6: Comparison of Bayesian Information Criteria for each Growth Mixture Model, all with eGFR as the main outcome (n=1827)

| Covariates | Number of classes | | | |
|---|---|---|---|---|
| | 1 | 2 | 3 | 4 |
| Sex | 15018.0 | 14989.0 | 14993.4 | 15006.9 |
| Sex + sex interaction with time | 15021.9 | 14993.0 | 14997.9 | 15012.1 |
| Sex and age | 14816.8 | 14696.7 | 14707.8 | 14722.9 |
| Sex + age + sex interaction with time | 14820.9 | 14701.8 | 14708.9 | 14722.7 |
| Sex, age + age-squared | 14822.8 | 14700.5 | 14706.0 | 14739.4 |
| Sex, age, age-squared + sex interaction with time | 14826.8 | 14705.6 | 14711.6 | 14735.6 |

Table S7: Growth Mixture Models selected model output (n=1827)

| Adjusting for | Class characteristics | | | |
|---|---|---|---|---|
| | Class/category | Class distribution (all; F; M) | Intercept (F; M) | Slope |
| Sex and age | 1 (high, stable) | 78.6%; 82.8%; 73.6% | 118.12; 116.53 | -1.10 |
| | 2 (lower, erratic) | 21.4%; 17.2%; 26.4% | 101.69; 100.10 | -1.83 |

Table S8: Category comparison between best Hidden Markov Model and best Growth Mixture Model

| Growth Mixture Model Class | Hidden Markov Model category | | | | |
|---|---|---|---|---|---|
| | Stayed healthy | Healthy to unhealthy | Stayed unhealthy | Reverted to healthy | Total |
| 1 (high stable eGFR) | 305 (91%) | 10 (3%) | 0 (0%) | 19 (6%) | 334 (100%) |
| 2 (unstable/lower eGFR) | 11 (12%) | 33 (36%) | 19 (21%) | 28 (31%) | 91 (100%) |
| Total | 316 (74%) | 43 (10%) | 19 (4%) | 47 (11%) | 425 (100%) |

Table S9: Associations between baseline exposure in study participants and probability of membership to the lower erratic group from the Growth Mixture Model using fractional logistic regression, adjusted for sex, age and education level.

<table>
<tr><th colspan="2" rowspan="2">Variable</th><th colspan="3">Association with lower, erratic group</th></tr>
<tr><th>n</th><th>OR (95% CI)</th><th>p-value</th></tr>
<tr><td colspan="2">Sex (Male)</td><td>425</td><td>1.50 (1.07, 2.10)</td><td>0.02</td></tr>
<tr><td colspan="2">Age (1-year increase)</td><td>425</td><td>1.01 (0.99, 1.03)</td><td>0.21</td></tr>
<tr><td rowspan="3">Education level</td><td>No O levels</td><td rowspan="3">425</td><td>1.10 (0.74, 1.61)</td><td>0.65</td></tr>
<tr><td>O levels</td><td>ref</td><td>NA</td></tr>
<tr><td>A levels and above</td><td>0.99 (0.60, 1.64)</td><td>0.97</td></tr>
<tr><td colspan="2">Main occupation is farming</td><td>425</td><td>1.14 (0.79, 1.63)</td><td>0.49</td></tr>
<tr><td rowspan="4">Duration of farming</td><td>No farming</td><td rowspan="4">420</td><td>ref</td><td>NA</td></tr>
<tr><td><10 years</td><td>0.90 (0.56, 1.44)</td><td>0.65</td></tr>
<tr><td><20 years</td><td>1.46 (0.87, 2.45)</td><td>0.16</td></tr>
<tr><td><30 years</td><td>1.25 (0.69, 2.24)</td><td>0.47</td></tr>
<tr><td rowspan="3">Income group</td><td>Low (<=20000)</td><td rowspan="3">425</td><td>1.18 (0.78, 1.79)</td><td>0.43</td></tr>
<tr><td>Medium</td><td>ref</td><td>NA</td></tr>
<tr><td>High (>40000)</td><td>0.96 (0.64, 1.45)</td><td>0.84</td></tr>
<tr><td rowspan="4">BMI category</td><td>Underweight</td><td rowspan="4">424</td><td>0.87 (0.55, 1.36)</td><td>0.54</td></tr>
<tr><td>Normal</td><td>ref</td><td>NA</td></tr>
<tr><td>Overweight</td><td>1.07 (0.70, 1.63)</td><td>0.77</td></tr>
<tr><td>Obese</td><td>1.02 (0.60, 1.71)</td><td>0.95</td></tr>
<tr><td colspan="2">Painkiller tablets</td><td>425</td><td>1.28 (0.86, 1.90)</td><td>0.23</td></tr>
<tr><td colspan="2">Painkiller injections</td><td>425</td><td>0.85 (0.34, 2.12)</td><td>0.73</td></tr>
<tr><td colspan="2">Current smoker</td><td>424</td><td>1.18 (0.70, 1.97)</td><td>0.54</td></tr>
<tr><td colspan="2">Current smokeless tobacco user</td><td>421</td><td>1.54 (0.96, 2.49)</td><td>0.08</td></tr>
<tr><td colspan="2">Drinks Alcohol 2+ times a week</td><td>410</td><td>1.74 (0.70, 4.31)</td><td>0.23</td></tr>
<tr><td colspan="2">Very physically active</td><td>422</td><td>1.14 (0.77, 1.67)</td><td>0.52</td></tr>
<tr><td colspan="2">Exposed exposure to sunlight</td><td>422</td><td>0.92 (0.64, 1.31)</td><td>0.64</td></tr>
<tr><td rowspan="3">Breaks from work when exposed</td><td>No sunlight</td><td rowspan="3">418</td><td>ref</td><td>NA</td></tr>
<tr><td>Sunlight with breaks</td><td>1.03 (0.72, 1.47)</td><td>0.98</td></tr>
<tr><td>Sunlight no breaks</td><td>0.63 (0.20, 2.02)</td><td>0.44</td></tr>
<tr><td rowspan="3">Breaks from work in shade when exposed</td><td>No sunlight</td><td rowspan="3">407</td><td>ref</td><td>NA</td></tr>
<tr><td>Breaks in shade</td><td>1.11 (0.74, 1.67)</td><td>0.61</td></tr>
<tr><td>No breaks in shade</td><td>0.94 (0.56, 1.59)</td><td>0.82</td></tr>
<tr><td colspan="2">Reverse Osmosis water use</td><td>422</td><td>1.33 (0.93, 1.90)</td><td>0.12</td></tr>
<tr><td colspan="2">Vegetarian diet</td><td>424</td><td>1.34 (0.50, 3.61)</td><td>0.56</td></tr>
<tr><td colspan="2">Fish in diet</td><td>425</td><td>0.60 (0.31, 1.17)</td><td>0.14</td></tr>
<tr><td colspan="2">Meat in diet</td><td>424</td><td>1.48 (0.88, 2.51)</td><td>0.14</td></tr>
<tr><td colspan="2">Eggs in diet</td><td>425</td><td>0.85 (0.47, 1.55)</td><td>0.61</td></tr>
<tr><td colspan="2">Fertilizer exposure</td><td>425</td><td>1.05 (0.71, 1.53)</td><td>0.82</td></tr>
<tr><td colspan="2">Weedkiller exposure</td><td>425</td><td>1.31 (0.87, 1.96)</td><td>0.19</td></tr>
<tr><td colspan="2">Roundup exposure</td><td>425</td><td>1.12 (0.70, 1.79)</td><td>0.63</td></tr>
<tr><td colspan="2">Pesticide exposure</td><td>425</td><td>1.15 (0.77, 1.71)</td><td>0.50</td></tr>
</table>

OR, odds ratio; 95% CI, 95% confidence interval; BMI, Body Mass Index
